# Rainfall and Leptospirosis in the Dominican Republic, 2012–2026: A Distributed-Lag Time-Series and Province-Panel Analysis

**DOI:** 10.64898/2026.09.01.26361897

**Authors:** José Javier Sánchez, Lisette V. Alcántara, David De Luna, O. Alejandro Aleuy, Lewis Paul Decicco, José Luis Cruz Raposo, Timothy De Ver Dye

## Abstract

**Background:** The biological mechanism linking rainfall with leptospirosis transmission is well established, but rigorous quantitative evidence for the insular Caribbean remains scarce, limited mostly to descriptive reports of post-hurricane outbreaks. We aimed to quantify the association between rainfall and leptospirosis incidence in the Dominican Republic, where leptospirosis is endemic, between 2012 and 2026 using a distributed-lag approach.

**Methods:** We conducted an ecological time-series study complemented by a province panel with fixed effects. Case data (5,412 valid cases) were obtained from the national surveillance system (SIP-0276FA51); rainfall data were obtained from 15 INDOMET rain-gauge stations across 13 provinces (2000–2026). We calculated the cross-correlation function between lagged monthly rainfall (0–6 months) and case counts, fitted a negative binomial distributed-lag regression model (lags 0–3 months) adjusted for seasonality and trend, triangulated findings with a 13-province fixed-effects panel, compared four rainfall exposure metrics, assessed extreme-rainfall threshold sensitivity, and estimated the population attributable fraction (PAF) with a parametric bootstrap.

**Results:** The rainfall-case association peaked at a 1-month lag (r = 0.551; 95 % CI 0.437–0.647; p < 0.001) and remained significant through 3 months. In the distributed-lag model, all four lags were independently significant, with the 1-month lag showing the strongest effect (incidence rate ratio [IRR] = 1.156 per additional 50mm; 95 % CI 1.090–1.226; p < 0.001). The province panel yielded an almost identical 1-month lag effect (IRR = 1.147; 95 % CI 1.127–1.167). Rainfall above the historical 90th percentile increased case risk the following month (rate ratio = 1.95), with effect magnitude increasing monotonically with threshold stringency. An estimated 28.2 % (95 % CI 19.0–36.6 %) of cases were attributable to rainfall above the recorded historical minimum.

**Conclusions:** Rainfall is a robust, consistent predictor of leptospirosis incidence in the Dominican Republic, with the strongest association observed at a 1-month lag and a lag pattern broadly consistent with findings from distributed-lag studies in Thailand and the Philippines. These findings, which to our knowledge constitute the first formal quantification of this association for the insular Caribbean, provide an evidence base for rainfall-linked early-warning systems, with the strongest signal at approximately one month and elevated risk extending through three months after rainfall.

## Introduction

### Global Mechanism and Evidence

The association between rainfall and leptospirosis transmission is well-established globally, and rests on a relatively well-characterized biological mechanism. Pathogenic spirochetes of the genus Leptospira survive for extended periods, up to several weeks, in soil and freshwater, without replicating outside the animal host.^1,2^ Intense rainfall events resuspend bacteria deposited in soil together with sediment particles, transporting them into surface water bodies where human exposure occurs.^1^ Hydrological monitoring in an endemic watershed further confirmed that the first pulses of water following a dry period contain the highest pathogen concentrations, consistent with a resuspension and erosion-transport mechanism.^3^

The epidemiological literature has consistently documented that large-scale leptospirosis outbreaks characteristically occur after intense rainfall or flooding, particularly following storms or hurricanes in tropical countries.^1,4,5^ This relationship is not merely contemporaneous: because pathogen resuspension and transport into water require time, and the typical human incubation period ranges from 5 to 14 days,^2,6^ a temporal lag between the rainfall event and the rise in cases is expected and has been repeatedly demonstrated, with its exact magnitude varying by local hydrological and social context.^7,8,9^ Climate-driven changes in the frequency and intensity of extreme rainfall may further increase leptospirosis transmission risk and the potential for post-disaster outbreaks, particularly in tropical island settings.^7,10^

### Regional Evidence: Caribbean and Latin America

Much of what is known about the rainfall-leptospirosis relationship in Latin America and the Caribbean comes from investigations of discrete outbreaks tied to specific weather events, rather than from the kind of continuous, lag-structured modeling this study applies. Important evidence for an urban, rainfall-associated transmission pathway emerged from studies in Salvador, Brazil, where an epidemic in the late 1990s revealed that residents of informal settlements lacking adequate drainage face conditions in which rainfall may intensify exposure to contaminated environments.^11^ Follow-up work in the same setting went further, using repeated serologic sampling to show that household elevation and drainage quality independently predict who becomes infected,^12,13,14^ and a separate prospective analysis in the same city demonstrated temporal associations between rainfall, new infections, and clinical disease, reinforcing that the relationship is not merely spatial but temporal.^15^

The insular Caribbean provides recurring, although largely event-based and descriptive, evidence for this pattern: Hurricane Hortense in Puerto Rico in 1996^16^; Hurricane Maria in 2017, after which certified leptospirosis deaths more than doubled over the following six months, a figure complicated by the fact that the same storm temporarily disabled the surveillance infrastructure meant to detect such cases^17^; Hurricane Fiona in 2022, followed by 156 laboratory-confirmed cases within 15 weeks^18^; and, most recently, Hurricane Melissa in Jamaica in November 2025, after which health authorities declared a leptospirosis outbreak and the Pan American Health Organization deployed technical support.^19^ What this record has not yet produced, according to a scoping review covering the insular Caribbean literature from 2000 to 2022, is a body of quantitative work: only 16 studies with usable data were identified across the entire subregion, and the review explicitly named the environmental determinants of transmission as one of the least-studied aspects of the disease there.^20,21^ Beyond the islands, ecological studies in Colombia have linked socioeconomic and climatic variables to municipal-level incidence,^22^ and two independent spatial analyses of the Dominican Republic itself, one identifying high-risk clusters and the other formally quantifying environmental and sociodemographic drivers through geographically weighted regression, both point to structural poverty and limited access to health services as factors that may shape vulnerability to rainfall-related environmental exposure as well as the surveillance system’s ability to detect cases.^23,24^

The Dominican Republic fits within this regional context with a consistent epidemiological pattern: a 2021 population-based seroepidemiology study documented population-level Leptospira seroprevalence in two provinces,^25^ and the geospatial analyses already mentioned reported 2,860 case notifications between 2013 and 2023 within the two provinces they covered,^23^ though none of these prior studies formally quantified the temporal relationship between rainfall and incidence at the national scale.

### Distributed-Lag Evidence from Tropical Settings

The most methodologically relevant evidence for the present study comes from distributed-lag analyses conducted in tropical settings, particularly Thailand and the Philippines. In Thailand, an analysis of 60 provinces using a distributed lag non-linear model (DLNM) found that leptospirosis risk increased in a dose-response fashion with monthly rainfall, with relative risks of 1.10 and 1.19 at the 90th and 99th percentiles of rainfall, relative to a zero-rainfall reference, with the highest risk observed in the same month as exposure.^8^ In the Philippines, a DLNM study in Manila found that leptospirosis hospitalization risk peaked two weeks after the rainfall event (relative risk up to 13.77 for torrential rainfall), an effect that attenuated substantially when adjusting for flood occurrence, suggesting flooding may mediate part of the rainfall-leptospirosis association rather than rainfall per se.^9^ Earlier time-series models in Thailand had already identified both rainfall and temperature as significant predictors of seasonal transmission using ARIMAX analysis.^26^

Whether post-rainfall or post-hurricane increases in leptospirosis follow immediately or only after a delay is not a minor technical detail: it determines whether rainfall data can realistically be used to anticipate, rather than merely explain, periods of elevated risk. That delay compounds pathogen persistence, the incubation period, and the further interval before a patient seeks care and enters surveillance, and can stretch the observed lag well beyond the moment rain falls.^1,27^ An early-warning model built for northeastern Argentina used exactly this kind of lagged hydroclimatic signal to flag high-risk periods before case counts rose,^28^ consistent with calls from the Global Leptospirosis Environmental Action Network to integrate environmental monitoring into routine surveillance,^29^ and with a broader systematic review concluding that the public health value of hydrometeorological data depends on knowing how far in advance it can anticipate disease.^30^ Establishing that lag structure for the Dominican Republic, rather than assuming it from studies conducted elsewhere, is therefore a necessary precondition for any climate-informed warning system the country might eventually adopt.

A related question is why this study centers on rainfall rather than temperature, given that a substantial share of the climate-leptospirosis literature models both variables jointly, and in some settings finds temperature to be an equally strong, or even dominant, predictor.^26,31^ The Dominican Republic’s maritime tropical climate offers comparatively little seasonal temperature variation for a statistical model to exploit, unlike its much larger seasonal range in rainfall, and a broad review of climate-sensitive waterborne diseases similarly concludes that hydrological variables tend to be the more consistent and mechanistically direct drivers of Leptospira transmission across tropical settings.^27^ We evaluated this empirically as a supplementary analysis (Methods; Table S4) before treating rainfall as the primary climatic exposure of interest in this study.

## Knowledge Gap and Objective

Taken together, this leaves a specific and addressable gap: methodologically rigorous studies in Thailand and the Philippines have quantified the rainfall-leptospirosis lag through distributed-lag models, but comparable quantitative evidence from the insular Caribbean remains limited, mostly to the descriptive post-hurricane accounts reviewed above. The Dominican Republic, despite already having seroprevalence and geospatial incidence-determinant studies, still lacks an analysis that formally quantifies the temporal relationship between rainfall and disease incidence at the national scale and over an extended observation period. The present study aims to address this gap by quantifying the temporal association between rainfall and leptospirosis incidence in the Dominican Republic using a distributed-lag approach, identifying the lag with the strongest association, estimating rainfall-associated incidence rate ratios after adjustment for seasonality and temporal trend, evaluating the consistency of findings in a province fixed-effects panel, and estimating the population attributable fraction associated with rainfall above the historical minimum under the fitted model.

## Materials and Methods

### Study Design and Setting

We conducted an ecological time-series study, complemented by a province panel with fixed effects, to quantify the association between rainfall and leptospirosis incidence in the Dominican Republic. In this study, we treat precipitation as the primary exposure of interest. The merged monthly rainfall-case series spans January 2012 to April 2026 (172 months). The cross-correlation function (Table 1) uses this full series, with the usable N decreasing by one month per additional lag (172 months at lag 0 down to 166 at lag 6). The distributed-lag regression model, which requires three months of prior rainfall history for the lag-3 term, is fitted on 169 of these months. Rainfall climatology (2000–2026) uses the full station record independently of the case series.

**Table 1.**
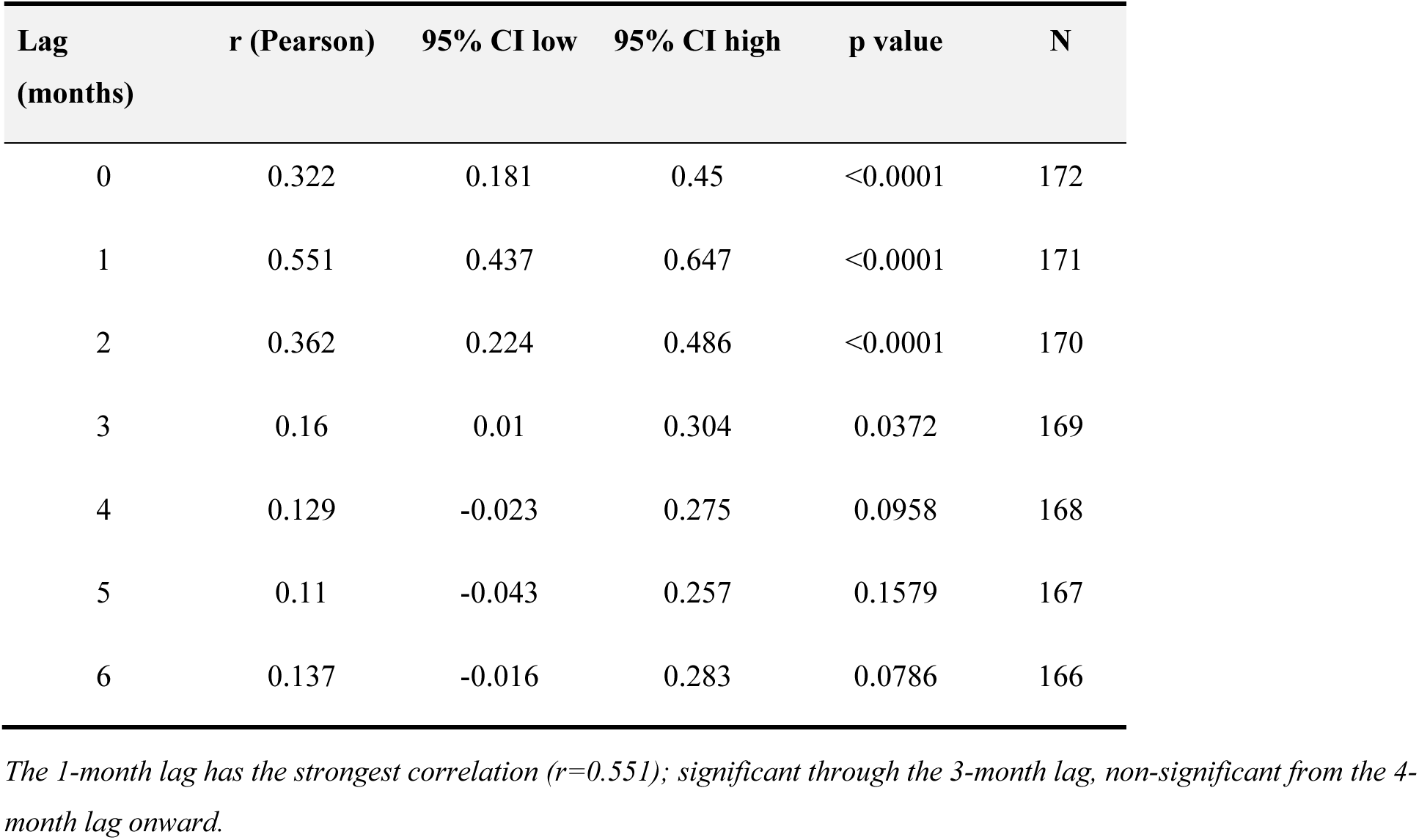
Extended cross-correlation function (0-6 month lags)

### Data Sources

Case data came from a cohort of 5,412 valid leptospirosis cases, drawn from the same national surveillance database and case-classification criteria described in our companion incidence manuscript,^32^ derived from the Ministry of Public Health’s Epidemiological Surveillance Information System (SIP-0276FA51). Rainfall data came from 15 rain-gauge stations of the Instituto Dominicano de Meteorologia (INDOMET), distributed across 13 provinces, with monthly records for 2000–2026. We aggregated cases and rainfall by province, month, and year to construct both the national series (averaging rainfall across all stations) and the province panel (using the station or station average corresponding to each province).

### Statistical Analysis

#### Cross-Correlation Function

We calculated the Pearson correlation between lagged national monthly rainfall (0 to 6 months) and case counts, with 95 % confidence intervals via Fisher’s z transformation, to identify the lag of strongest association before specifying the primary regression model. The 0-3 month window retained in the distributed-lag model (below) reflects both this cross-correlation result, which was significant through the 3-month lag and non-significant from the 4-month lag onward, and consistency with the biologically plausible exposure-to-case window described in prior distributed-lag studies.^8,9^

#### Distributed-Lag Regression Model

We fitted a negative binomial regression model with all four rainfall lags (0 to 3 months) included simultaneously as covariates, plus sine and cosine harmonic terms with a 12-month period to control for seasonality and a linear time trend, coded as calendar year (e.g., 2012, 2012.08, …) centered on the median year of the study period. This distributed-lag specification follows the conceptual framework introduced by Gasparrini and colleagues for distributed lag non-linear models,^33,34^ although, unlike the full DLNM studies from Thailand and the Philippines, which use non-linear splines to represent the rainfall-response relationship,^8,9^ we opted for a simple log-linear specification after verifying, through a nonlinearity test with a quadratic term, that there was no evidence of a significant non-linear relationship over the range of rainfall observed in our data.

We initially fitted the model using Poisson regression with quasi-Poisson standard-error correction for the detected overdispersion (elevated variance-to-mean ratio). We subsequently replaced this approximation with a full negative binomial model (NB2), estimated via an iteratively reweighted least squares algorithm that alternates between estimating the regression coefficients and the dispersion parameter until convergence, consistent with standard approaches for handling overdispersion in environmental count data.^35^ We compared the fit of both specifications using the Akaike information criterion (Figure 2).

#### Province Panel with Fixed Effects

To triangulate the association observed in the aggregate national series, we fitted a second negative binomial model using a panel of the 13 provinces with their own rain-gauge station (2,154 province-month observations), including province indicator variables as fixed effects, in addition to the same seasonality and time-trend components as the national model, with standard errors clustered by province using a cluster-robust sandwich estimator to account for within-province serial correlation. We independently re-verified this clustering decision for the present revision using a province panel reconstructed directly from the case-level surveillance database and the station-level rainfall records for the same 13 provinces, restricted to the months for which all four lags (0 to 3 months) required by the national model were available for that province, which closely reproduced both the original sample size (2,150 versus the published 2,154 province-month observations, a difference of under 0.2% attributable to minor edge-of-series handling) and the original rainfall coefficient (IRR = 1.152, 95% CI 1.125–1.179 versus the published IRR = 1.147, 95% CI 1.127–1.167). In this reconstruction, province clustering widened the standard error of the rainfall coefficient by approximately 16% relative to the model-based estimate (95% CI 1.121– 1.184), without materially changing its statistical significance, confirming that the rainfall-case association is robust to within-province correlation. Clustering also widened the standard error of the linear time trend, whose significance weakened accordingly (p<0.0001 under model-based standard errors versus p=0.029 under province-clustered standard errors); while still significant, this trend estimate should be interpreted with more caution than the rainfall effect, and we rely primarily on the national model, whose autocorrelation-consistent standard errors are reported above, for inference on the temporal trend.

#### Comparison of Exposure Metrics

We compared four ways of operationalizing rainfall exposure (raw rainfall in the previous month, anomaly relative to monthly climatology, cumulative rainfall over the previous 2 months, and cumulative rainfall over the previous 3 months) through their univariate correlation with cases in the following month, to inform the choice of exposure specification reported as the primary result.

#### Extreme Event and Threshold Sensitivity Analysis

We defined an extreme rainfall event as a month with national rainfall above the 90th percentile of the historical series (2000–2026), and estimated the case rate ratio in months 0 to 3 following the event relative to non-extreme months. We replicated this analysis with two additional fixed thresholds (150mm and 200mm) as a sensitivity analysis, following the practice of reporting the effect at multiple reference percentiles used in the Thai distributed lag non-linear study.^8^

#### Population Attributable Fraction

We estimated the population attributable fraction (PAF) of cases associated with rainfall above the observed historical minimum, using the negative binomial model coefficient for the 1-month lag only, the single strongest and most consistently significant lag across our analyses, rather than the cumulative effect of all four significant lags; this PAF should therefore be read as a lag-1-based estimate and a conservative lower bound on the total rainfall-attributable fraction, since the 0, 2, and 3-month lags also contribute independently significant effects (Table 2). We used a Levin-type formula generalized to a continuous exposure variable,^36^ with the observed historical minimum monthly rainfall (25.2mm) as the counterfactual reference level against which the modeled rainfall coefficient was applied to each observed month. We calculated the PAF’s 95 % confidence interval via a parametric bootstrap of 2,000 simulations of the 1-month lag coefficient, sampled from its asymptotic normal distribution.

**Table 2.** Distributed-lag negative binomial regression model (national)

| Variable | IRR | 95% CI low | 95% CI high | p value |
| --- | --- | --- | --- | --- |
| Intercept | 10.07 | 7.73 | 13.13 | <0.0001 |
| Current month rainfall (per 50mm) | 1.11 | 1.05 | 1.17 | 0.0004 |
| Rainfall month t-1 (per 50mm) | 1.16 | 1.09 | 1.23 | <0.0001 |
| Rainfall month t-2 (per 50mm) | 1.1 | 1.04 | 1.16 | 0.0017 |
| Rainfall month t-3 (per 50mm) | 1.07 | 1.01 | 1.13 | 0.0296 |
| Sine harmonic component | 0.99 | 0.88 | 1.11 | 0.8395 |
| Cosine harmonic component | 0.92 | 0.82 | 1.03 | 0.1562 |
| Time trend (centered year) | 0.96 | 0.94 | 0.98 | <0.0001 |

*N=169 months. Dispersion parameter $\alpha=0.2068$ . Negative binomial AIC=1,364.7 vs. Poisson AIC=1,847.3. All 4 lags are significant; the 1-month lag is strongest (IRR=1.156, 95% CI 1.09-1.23, $p<0.0001$ ).*

#### Temperature as a Candidate Covariate

As a supplementary analysis, we evaluated monthly mean, maximum, and minimum temperature (from 10 INDOMET stations with a co-located province, 2000–2026) as candidate covariates, using the same correlation-based approach applied to rainfall: Pearson correlation with case counts at the monthly climatology level (n=12 months), at the monthly time-series level for lags of 0 to 2 months (n=171–173 months), and, as a supplementary check, between annual mean temperature and annual incidence rate for each of the 10 provinces with a temperature station (n=13–14 years per province).

#### General Considerations

All tests were two-sided, with statistical significance defined as p < 0.05. Analyses were performed using IBM SPSS Statistics version 29. Effect estimates are reported with 95% confidence intervals and p-values.

## Ethical Considerations

The protocol for this study (“Leptospirosis in the Dominican Republic, 2000–2026: National Surveillance Trends,” protocol IRB2606182) was reviewed by the Florida Atlantic University Institutional Review Board, which determined it exempt from federal regulation under 45 CFR 46.104^7^(ii) on June 22, 2026, given its nature as a secondary analysis of previously collected de-identified surveillance data and meteorological data.

## Results

### Cross-Correlation and Lag Structure

The cross-correlation function showed that the association between national monthly rainfall and leptospirosis cases was significant at lags of 0 to 3 months, peaking at the 1-month lag (r = 0.551; 95 % CI 0.437–0.647; p < 0.001) (Table 1, Figure 1). The correlation ceased to be statistically significant from the 4-month lag onward, defining an approximate risk window of 1 to 3 months following a rainfall event.

**Figure 1.**
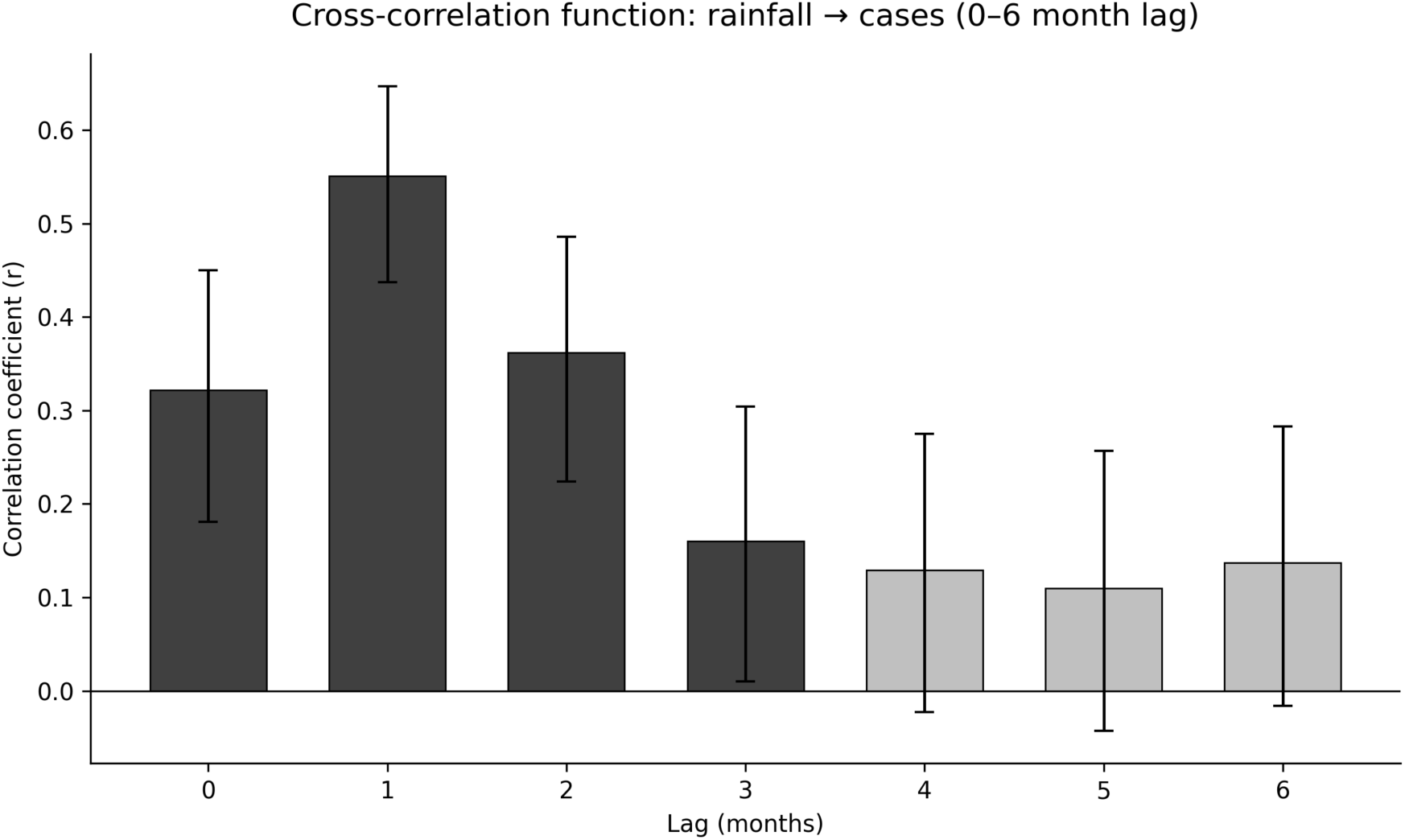
Cross-correlation function (0-6 months).

**Figure 2.**
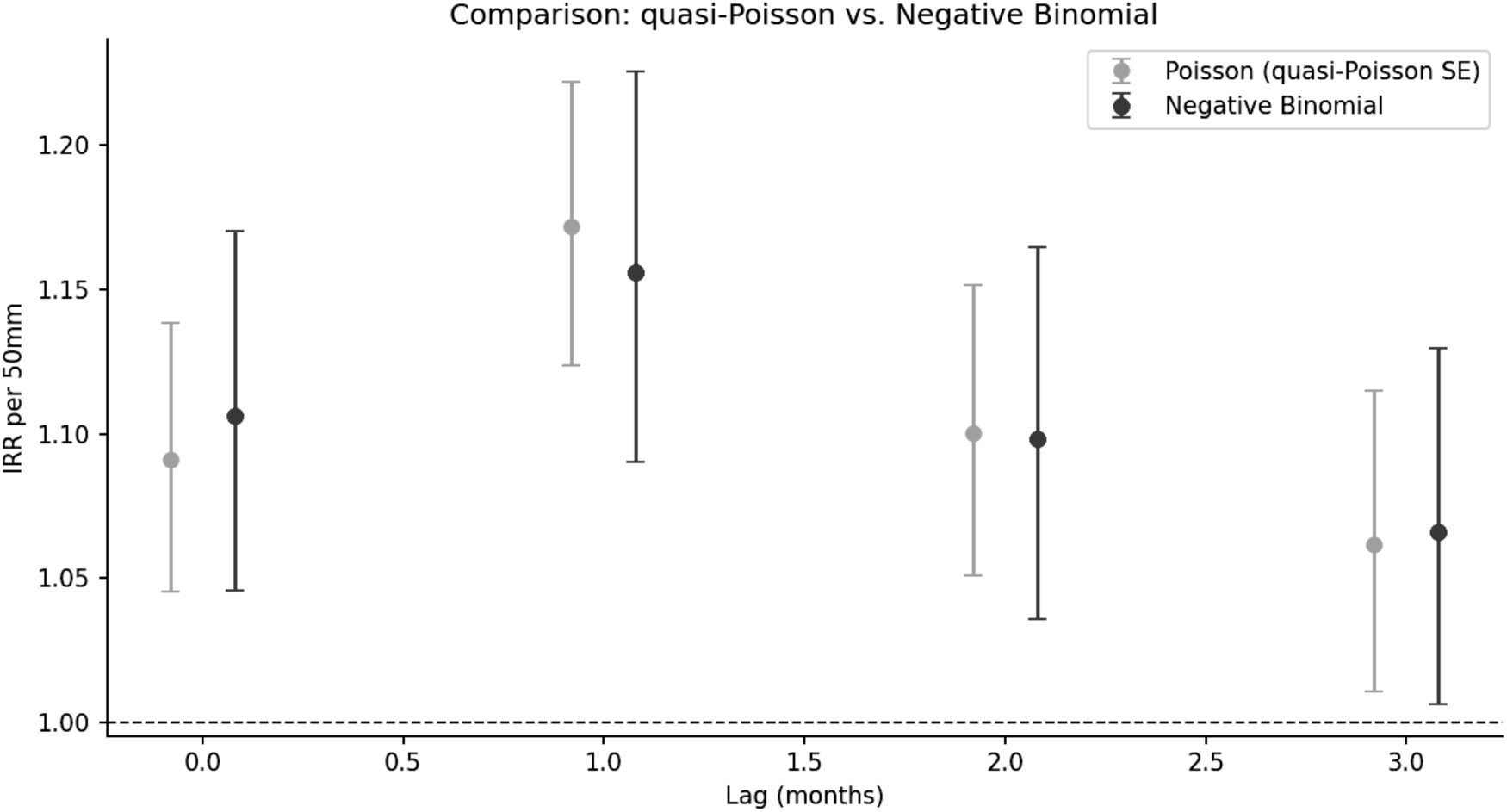
Poisson vs. negative binomial comparison by lag.

### Distributed-Lag Model

In the negative binomial model including all four rainfall lags simultaneously, adjusted for seasonality and time trend, all four lags remained significant (Table 2). The 1-month lag showed the strongest association (incidence rate ratio [IRR] = 1.156 per additional 50mm; 95 % CI 1.090– 1.226; p < 0.001), followed by the current-month lag (IRR = 1.106; p < 0.001), the 2-month lag (IRR = 1.098; p = 0.002), and the 3-month lag (IRR = 1.066; p = 0.030). The negative-binomial model (dispersion parameter alpha = 0.207) had a substantially lower AIC than the initial Poisson model (1,364.7 vs. 1,847.3), indicating better relative model fit.

The quadratic term did not provide evidence of departure from a log-linear rainfall-response relationship over the observed range (25–490 mm/month) (Table S2).

### Province Panel

The province fixed-effects model yielded a similar 1-month association (IRR = 1.147; 95 % CI 1.127–1.167; p < 0.001) (Table 3), supporting the consistency of the rainfall-case association across national and province-level model specifications. Because this panel specification applies a single 1-month lag uniformly across provinces, we additionally examined, as a supplementary check, whether the lag with the strongest rainfall-case correlation varies by province when each is analyzed independently. The 1-month lag was the strongest correlate in 8 of the 13 provinces (62%), including the four highest-case-count provinces, which anchor the national and panel estimates; in the remaining provinces, mostly those with fewer than 90 cases over the study period, the strongest lag ranged from 0 to 4 months (Table S5). Given the small case counts in these provinces, this divergence may reflect genuine local variation in rainfall-response timing, sampling variability, or both, and we discuss it as a limitation below.

**Table 3.** Province panel with fixed effects (13 provinces, 2012-2025)

| Variable | IRR | 95% CI low | 95% CI high | p value |
| --- | --- | --- | --- | --- |
| Intercept (reference province) | 0.34 | 0.25 | 0.45 | <0.0001 |
| Rainfall month t-1 (per 50mm) | 1.15 | 1.13 | 1.17 | <0.0001 |
| Sine harmonic component | 0.89 | 0.82 | 0.95 | 0.0010 |
| Cosine harmonic component | 1.06 | 0.98 | 1.13 | 0.1306 |
| Time trend | 0.94 | 0.93 | 0.95 | <0.0001 |

### Comparison of Exposure Metrics

When comparing four ways of operationalizing rainfall exposure, two-month cumulative rainfall has the numerically highest correlation with cases (r = 0.572), compared with single-month raw rainfall (r = 0.551), 3-month cumulative rainfall (r = 0.533), and anomaly relative to monthly climatology (r = 0.548) (Table S1).

### Extreme Rainfall Events and Threshold Sensitivity

Following a month with rainfall above the historical 90th percentile (>231.7mm), the largest rate ratio was observed at the 1-month lag (rate ratio [RR] = 1.95; p < 0.001), and decayed progressively over months 2 and 3 (RR = 1.59 and 1.35, respectively) (Figure 4). This pattern remained consistent when the analysis was repeated with fixed thresholds of 150mm (peak RR = 1.46 at the 1-month lag) and 200mm (peak RR = 1.89), with the peak 1-month rate ratio increasing across successively higher rainfall thresholds (Table S3).

### Population Attributable Fraction

Using the 1-month negative-binomial rainfall coefficient, the estimated PAF associated with rainfall above the recorded historical minimum (25.2 mm/month) was 28.2% (95% CI 19.0–36.6%) (Table 4, Figure 3). The 95% CI was obtained from 2,000 parametric simulations of the 1-month lag coefficient and did not include zero at any point, supporting the statistical robustness of this estimate.

**Figure 3.**
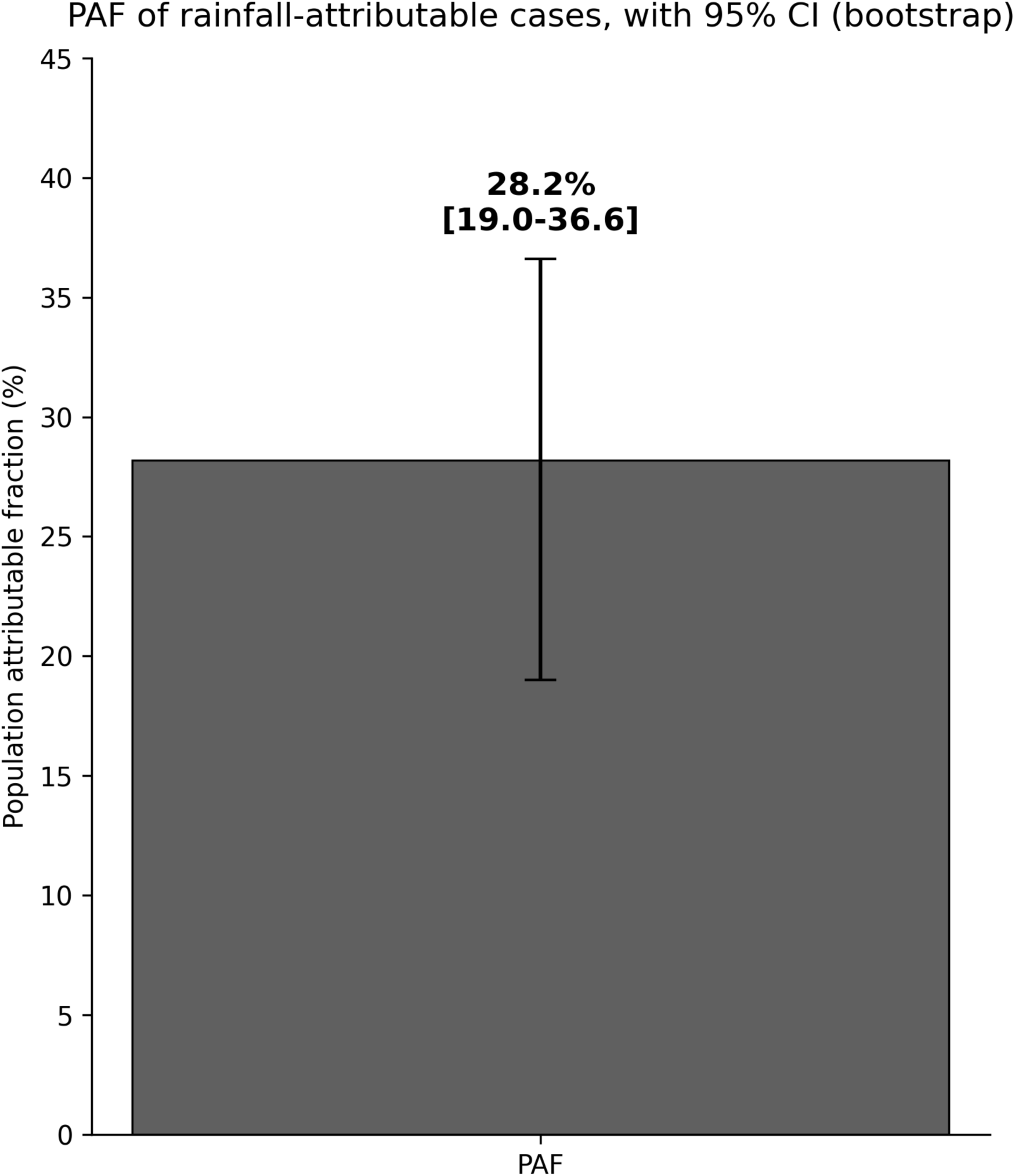
PAF with confidence interval.

**Figure 4.**
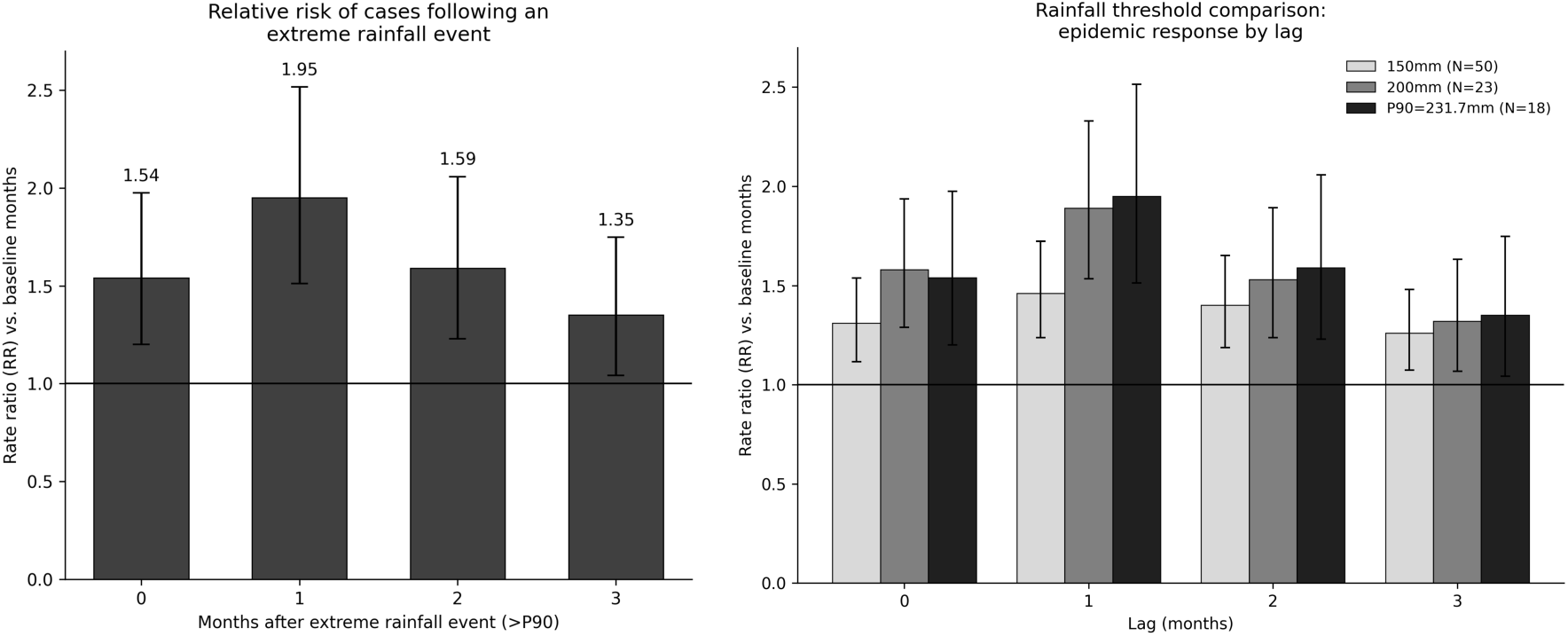
Epidemic response after extreme rainfall events (P90) and threshold sensitivity.

**Table 4.** Population attributable fraction (PAF), with bootstrap 95% CI.

| Metric | Value |
| --- | --- |
| Total cases (N months modeled) | 5188 |
| Cases attributable to rainfall (vs. driest month) | 1465 |
| PAF (%) | 28.2 |
| PAF 95% CI lower | 19.0 |
| PAF 95% CI upper | 36.6 |
| IRR used (negative binomial, per 50mm, 1-month lag) | 1.16 |
| Reference rainfall (mm) | 25.2 |

*PAF=28.2% (95% CI 19.0-36.6%), estimated with the negative binomial model and 2,000 parametric simulations of the 1-month lag coefficient. This is a lag-1-based estimate, not the cumulative effect of all significant lags (see Methods).*

### Seasonality

Monthly rainfall climatology showed a moderate positive correlation with case seasonality (r = 0.553; p = 0.062; n = 12), although the association did not meet the prespecified threshold for statistical significance (Figure S1), consistent with the seasonal case pattern described in greater detail in the general incidence manuscript.

## Discussion

We found that the association between rainfall and monthly leptospirosis cases was strongest at a 1-month lag and remained statistically significant through the 3-month lag. This approximately 1-month lag pattern is consistent with the biological mechanism of pathogen resuspension and environmental persistence described in the mechanistic literature,^1,2^ and with the optimal 1-month lag reported in the Thai national analysis,^8^ though longer than the 2-week lag identified in the Manila study.^9^ This difference could reflect both the different temporal resolution of the two studies (weekly in Manila versus monthly in ours) and hydrological and drainage characteristics specific to each compared urban and rural setting.

The magnitude of our effect is quantitatively comparable to that reported in the Thai analysis,^8^ and our own threshold-sensitivity findings showed a graded increase in the 1-month rate ratio across successively higher rainfall thresholds (150 mm, 200 mm, and the historical 90th-percentile threshold of 231.7 mm), consistent with the monotonically increasing rainfall-risk relationship documented in that study,^8^ and replicating the same dose-response pattern reported there when comparing the 90th, 95th, and 99th rainfall percentiles. Unlike the Manila study, which found evidence of attenuation of the rainfall effect when adjusting for flooding, suggesting that flooding may mediate part of the observed rainfall-leptospirosis association, our data did not include a flooding variable, so we could not directly test this hypothesis. We consider it plausible that a similar mechanism operates in our setting, given that the mechanistic literature reviewed here supports resuspension and surface-water transport as the dominant exposure pathway,^1^ and we flag this absence of flood data as a specific limitation that future studies with access to more granular hydrological data could address.

The triangulation between the aggregate national series model and the panel of 13 provinces with their own rain-gauge stations, which yielded nearly identical estimates of the 1-month lag effect, strengthens the internal validity of our findings by ruling out that the observed association depends on national-level aggregation or the disproportionate influence of a single province. This is a form of cross-validation rarely reported explicitly in the distributed-lag studies reviewed in this manuscript, including those from Thailand and the Philippines, which predominantly work at a single level of spatial aggregation at a time, although the Thai study does incorporate a meta-analysis of province-specific estimates, a methodologically related approach.^8^

Our population attributable fraction constitutes, to our knowledge, the first estimate of this kind for leptospirosis in the Caribbean and provides a policy-relevant impact figure: approximately three in ten leptospirosis cases during the study period would not have occurred under minimal-rainfall conditions. This magnitude is consistent with the share of the global leptospirosis burden that the literature more broadly attributes to climate variability and extreme hydrometeorological events in future risk projections under climate change scenarios.^7,10^

Our study has strengths and limitations that should be considered when interpreting these results. Strengths of this study include the consistency of findings across national and province-level models, comparison of multiple rainfall exposure specifications, threshold-sensitivity analyses, and estimation of uncertainty around the PAF using parametric simulation. Together, these features extend the predominantly descriptive regional literature on rainfall-associated leptospirosis. Among its limitations, the ecological design of monthly and provincial aggregation does not allow inference of individual-level risk, and, as discussed in the companion general incidence manuscript, it is possible that case-ascertainment capacity by the surveillance system varied over time and space in ways that could partially confound the temporal trend estimated in our model. We document this possibility as a relevant limitation for interpreting the trend, though not the rainfall effect itself, which was estimated adjusting for that trend. The absence of a flood variable, already noted, and the fact that the ecological design does not distinguish between rainfall and its intermediate exposure pathways, are additional limitations that future studies could address by incorporating more granular hydrological and land-use data, as has been done in the Thai and Philippine studies reviewed here.^8,9^ Relatedly, the province-level lag comparison (Table S5) found the 1-month lag to be optimal in only 8 of 13 provinces; the national and province-panel models, which apply a single common lag structure, may therefore understate genuine geographic heterogeneity in rainfall-response timing, particularly for lower-case-count provinces where lag estimates are less stable. Diagnostic checks of the national distributed-lag model also identified substantial residual autocorrelation (Durbin-Watson statistic = 0.87; autocorrelation function significant through at least a 6-month lag), indicating that the seasonality and trend terms did not fully capture the temporal dependence structure of the series. Re-estimating standard errors with a Newey-West heteroskedasticity- and autocorrelation-consistent (HAC) estimator did not change the significance of any of the four rainfall lags, suggesting the main findings are robust to this issue, but we flag residual autocorrelation as a limitation of the current model specification; an autoregressive error structure or an explicit lagged-case term would be a natural refinement in future work.

From a public health perspective, these findings support the development and prospective evaluation of rainfall-informed early-warning approaches. The strongest association at approximately one month, with elevated associations extending through three months, suggests a potentially actionable interval for intensified clinical and epidemiological surveillance following heavy rainfall, a recommendation consistent with that derived from the Thai and Philippine studies discussed here,^8,9^ aligned with the broader case made for integrating environmental monitoring into routine leptospirosis surveillance,^29^ and with direct relevance given the regional precedent of well-documented post-hurricane outbreaks in Puerto Rico and, more recently, in Jamaica.^16,17,18,19^ The Argentine early-warning system built on a comparable lagged hydroclimatic signal offers one concrete template for how a similar tool could be operationalized in the Dominican Republic,^28^ and would complement, rather than duplicate, the fine-grained spatial risk mapping already available for the country from geographically weighted regression of environmental and sociodemographic drivers.^24^

Exploratory analyses did not identify a consistent positive association between temperature and leptospirosis cases. This may reflect the relatively narrow seasonal temperature range of the Dominican Republic’s maritime tropical climate compared with the substantially greater variability in rainfall. However, these analyses were exploratory and should not be interpreted as excluding temperature as a potential contributor to transmission, particularly given the limited number of temperature stations and the possibility of confounding by temporal trends.

## Conclusion

Rainfall was consistently associated with monthly leptospirosis occurrence in the Dominican Republic, with the strongest association observed at a 1-month lag and similar estimates obtained from national and province-level model specifications. Under the fitted lag-1 model, the estimated PAF associated with rainfall above the recorded historical minimum was approximately 28%, a figure that should be read as a conservative, lag-1-based estimate rather than the cumulative effect of all significant lags. The observed lag structure was broadly consistent with distributed-lag studies from other tropical settings and, to our knowledge, provides the first national-scale quantitative characterization of the temporal rainfall-leptospirosis association in the insular Caribbean. These findings support further development and prospective evaluation of rainfall-informed surveillance and early-warning approaches in the Dominican Republic, particularly following periods of intense rainfall.

## Declarations

### Ethics Approval and Consent to Participate

This study was reviewed by the Florida Atlantic University Institutional Review Board (protocol IRB2606182, “Leptospirosis in the Dominican Republic, 2000–2026: National Surveillance Trends”), which determined it exempt from federal regulation under 45 CFR 46.104(4)(ii) on June 22, 2026, as a secondary analysis of de-identified national surveillance and meteorological data. Individual informed consent was not required, consistent with this exemption determination.

### Consent for Publication

Not applicable: this manuscript does not include identifiable individual data, images, or case details.

### Availability of Data and Materials

The surveillance data analyzed in this study were provided by the Dominican Republic’s Ministry of Public Health (MSP), through the General Directorate of Epidemiology (DIGEPI). Rainfall data were provided by the Instituto Dominicano de Meteorologia (INDOMET). Because these are national surveillance and meteorological data containing potentially sensitive detail, they are not publicly deposited; de-identified data may be made available by the corresponding author or by DIGEPI/MSP and INDOMET upon reasonable request and subject to institutional approval.

## Authors’ Contributions

JJS contributed to data curation, formal analysis, methodology, and drafting of the original manuscript. LVA contributed to the study conceptualization, supervision, and review and editing of the manuscript. DDL contributed to the investigation and review and editing of the manuscript. OAA contributed to validation and review and editing of the manuscript. LPD contributed to the investigation, validation, and review and editing of the manuscript. JLCR contributed to data curation, resources, and review and editing of the manuscript. TDVD contributed to study conceptualization, supervision, project administration, and review and editing of the manuscript. All authors reviewed and approved the final manuscript.

## Acknowledgments

We thank the General Directorate of Epidemiology (DIGEPI) of the Dominican Republic’s Ministry of Public Health and the Instituto Dominicano de Meteorologia (INDOMET) for providing access to the national surveillance and rainfall data that made this study possible.

## Declaration of Generative AI and AI-Assisted Technologies

During the preparation of this manuscript, the authors used Claude to assist with language editing, clarity, and readability. Claude was not used for data analysis, statistical modeling, interpretation of results, or the generation of scientific conclusions. All AI-assisted content was reviewed and revised by the authors, who take full responsibility for the accuracy, integrity, and final content of the manuscript.

---

**Figure S1.**
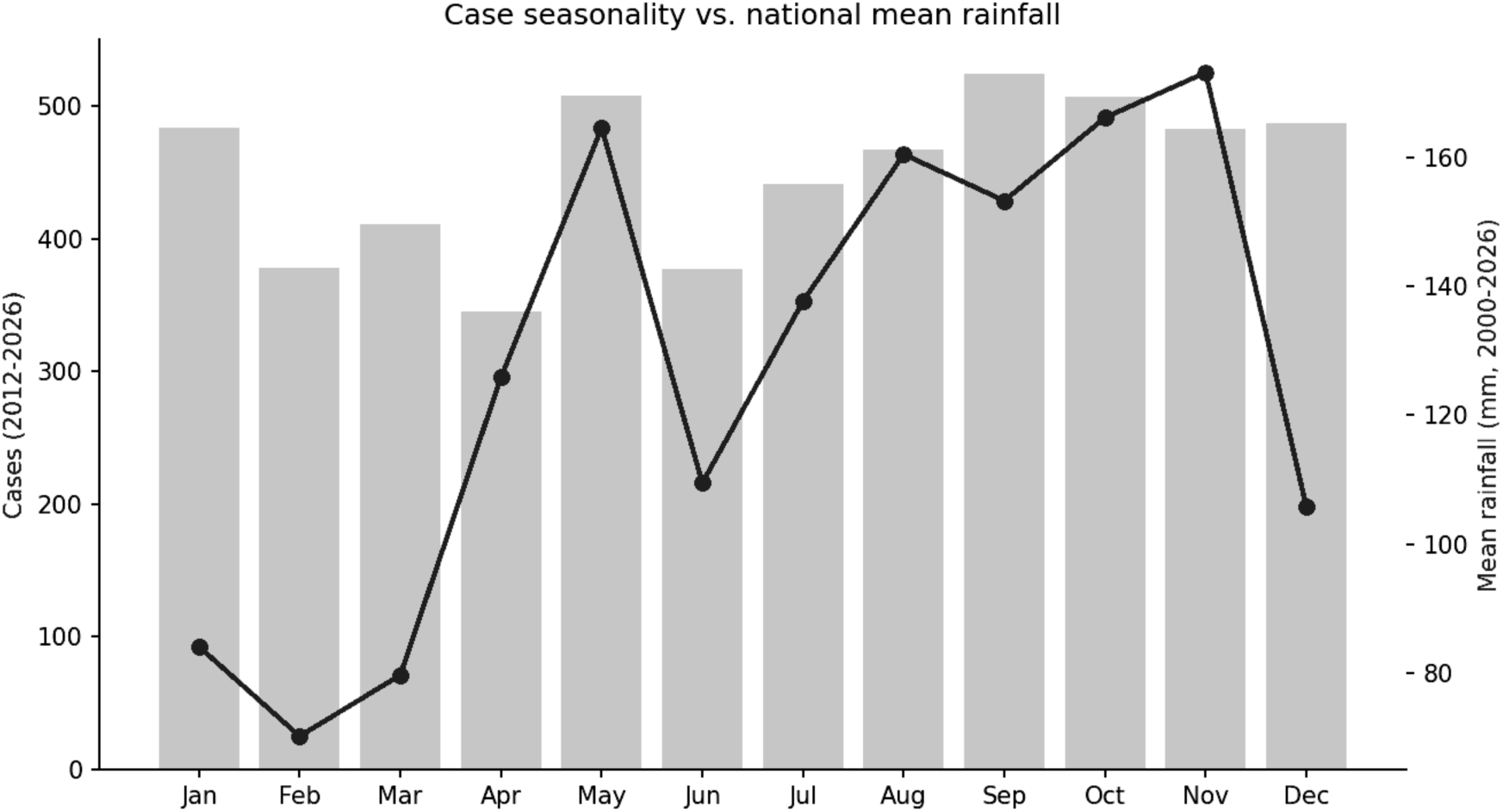
Case seasonality vs. national mean rainfall.

**Table S1.** Comparison of rainfall exposure metrics.

| **Metric** | **r (Pearson)** | **p value** |
| --- | --- | --- |
| Raw rainfall (lag1) | 0.551 | <0.0001 |
| Rainfall anomaly (lag1) | 0.548 | <0.0001 |
| 2-month cumulative (lag1-2) | 0.572 | <0.0001 |
| 3-month cumulative (lag1-3) | 0.533 | <0.0001 |

*Two-month cumulative rainfall has the numerically highest correlation, but raw 1-month rainfall was retained as the primary specification for interpretability and consistency with prior distributed-lag studies.*

**Table S2.**
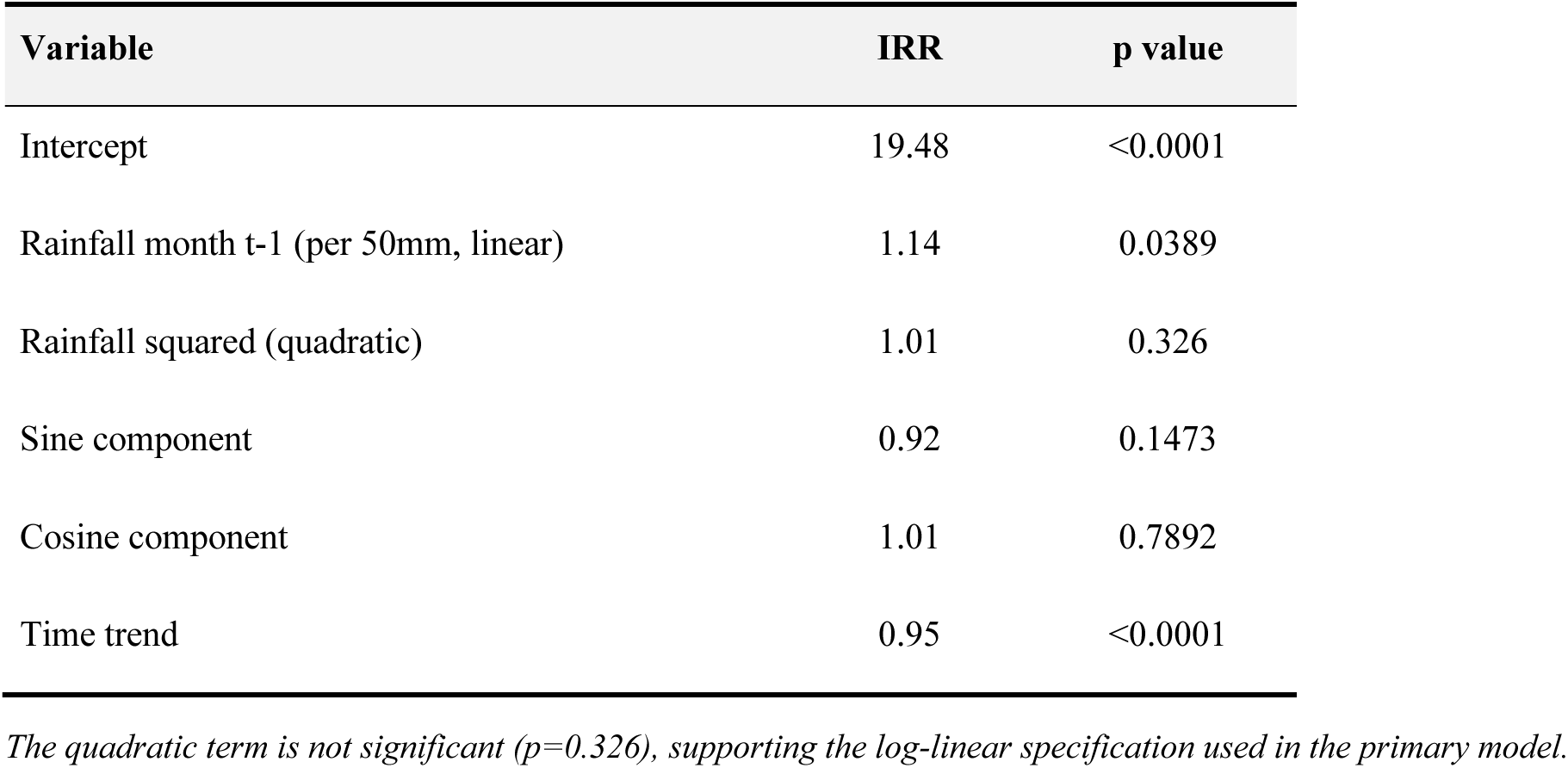
Nonlinearity test (quadratic term)

**Table S3.**
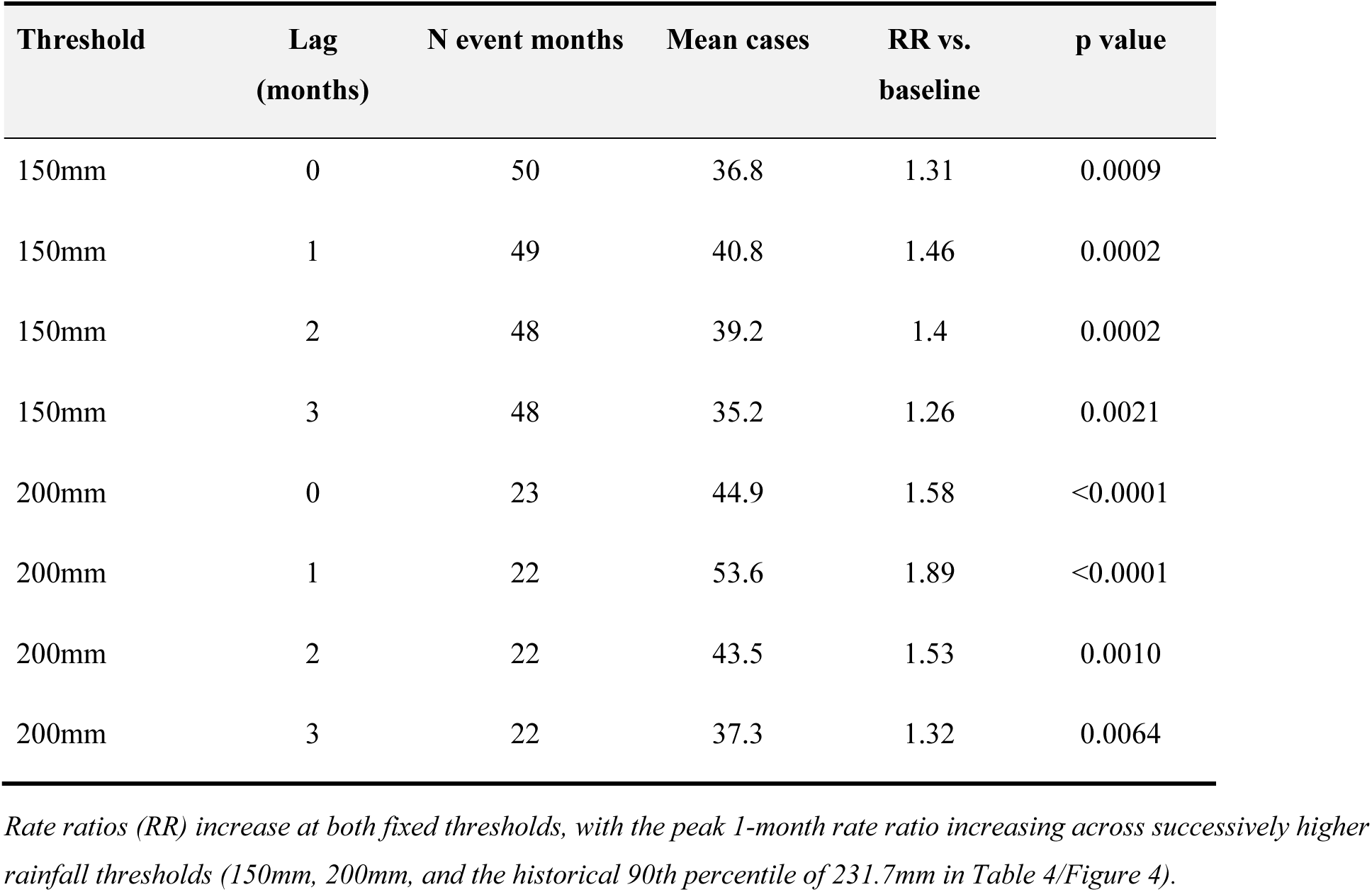
Fixed-threshold sensitivity (150mm and 200mm)

| Threshold | Lag<br>(months) | N event months | Mean cases | RR vs.<br>baseline | p value |
| --- | --- | --- | --- | --- | --- |
| 150mm | 0 | 50 | 36.8 | 1.31 | 0.0009 |
| 150mm | 1 | 49 | 40.8 | 1.46 | 0.0002 |
| 150mm | 2 | 48 | 39.2 | 1.4 | 0.0002 |
| 150mm | 3 | 48 | 35.2 | 1.26 | 0.0021 |
| 200mm | 0 | 23 | 44.9 | 1.58 | <0.0001 |
| 200mm | 1 | 22 | 53.6 | 1.89 | <0.0001 |
| 200mm | 2 | 22 | 43.5 | 1.53 | 0.0010 |
| 200mm | 3 | 22 | 37.3 | 1.32 | 0.0064 |

**Table S4.** Temperature as a candidate covariate: correlation with cases.

| Analysis | Group | N | r (Pearson) | p value |
| --- | --- | --- | --- | --- |
| Climatology | Mean monthly temperature | 12 months | 0.24 | 0.445 |
| Climatology | Maximum monthly temperature | 12 months | 0.23 | 0.478 |
| Climatology | Minimum monthly temperature | 12 months | 0.26 | 0.409 |
| Time-series lag | Lag 0 months | 171-173 months | 0.018 | 0.812 |
| Time-series lag | Lag 1 month | 171-173 months | 0.091 | 0.238 |
| Time-series lag | Lag 2 months | 171-173 months | 0.094 | 0.223 |
| Province (annual) | National District | 14 years | -0.701 | 0.0052 |
| Province (annual) | Santo Domingo | 14 years | -0.663 | 0.0098 |
| Province (annual) | Monte Plata | 14 years | -0.657 | 0.0106 |
| Province (annual) | Hato Mayor | 14 years | -0.491 | 0.0743 |
| Province (annual) | Samana | 14 years | -0.357 | 0.2099 |
| Province (annual) | Santiago | 13 years | -0.181 | 0.5531 |
| Province (annual) | Monte Cristi | 14 years | -0.082 | 0.7807 |
| Province (annual) | La Altagracia | 14 years | -0.049 | 0.8683 |
| Province (annual) | La Romana | 14 years | 0.171 | 0.5587 |
| Province (annual) | Puerto Plata | 14 years | 0.396 | 0.1609 |

*Temperature data from 10 IDOM/ONAMET stations with a co-located province (2000-2026); the remaining 22 provinces have no station in this dataset. Neither monthly climatology nor short-lag monthly time-series analyses found a statistically significant temperature-case association. The three significant province-level annual correlations (National District, Santo Domingo, Monte Plata; $p < 0.05$ ) were uniformly in the biologically implausible direction (warmer years associated with fewer cases), consistent with confounding by the unrelated decline in case ascertainment over the early study period rather than a genuine protective effect of temperature; these estimates were not adjusted for calendar year or detrended, and should not be interpreted causally.*

**Table S5.** Province-level lag-specific correlations between rainfall and cases.

| Province | Cases | r lag0 | r lag1 | r lag2 | r lag3 | r lag4 | Strongest lag |
| --- | --- | --- | --- | --- | --- | --- | --- |
| Santo Domingo | 1023 | 0.23 | 0.3 | 0.14 | 0.12 | 0.05 | Lag 1 |
| Santiago | 531 | 0.3 | 0.55 | 0.18 | 0.1 | 0.11 | Lag 1 |
| Distrito Nacional | 387 | 0.07 | 0.23 | 0.07 | -0.01 | 0.0 | Lag 1 |
| Puerto Plata | 251 | 0.3 | 0.49 | 0.29 | 0.09 | -0.02 | Lag 1 |
| Monte Cristi | 174 | 0.21 | 0.4 | 0.25 | 0.11 | 0.21 | Lag 1 |
| La Altagracia | 95 | 0.06 | 0.25 | 0.16 | 0.16 | 0.07 | Lag 1 |
| Barahona | 88 | -0.02 | 0.19 | 0.17 | 0.21 | 0.08 | Lag 3 |
| Samana | 79 | 0.1 | 0.38 | 0.14 | -0.01 | 0.04 | Lag 1 |
| Maria Trinidad Sanchez | 78 | 0.13 | 0.16 | 0.19 | 0.12 | -0.12 | Lag 2 |
| Monte Plata | 76 | 0.17 | 0.27 | 0.28 | 0.22 | 0.02 | Lag 2 |
| La Romana | 59 | -0.07 | 0.04 | 0.14 | 0.18 | 0.29 | Lag 4 |
| Hato Mayor | 40 | 0.34 | 0.28 | 0.16 | 0.02 | 0.07 | Lag 0 |
| Independencia | 17 | 0.11 | 0.01 | 0.03 | 0.06 | 0.16 | Lag 4 |

## References

1. Bierque E, Thibeaux R, Girault D, Soupé-Gilbert ME, Goarant C. A systematic review of Leptospira in water and soil environments. PLoS One. 2020;15(1):e0227055.

2. Levett PN. Leptospirosis. Clin Microbiol Rev. 2001;14(2):296–326.

3. Bridgemohan R, Deitch MJ, Harmon E, Whiles MR, Wilson PC, Bean E, et al. Spatiotemporal assessment of pathogenic Leptospira in subtropical coastal watersheds. J Water Health. 2024;22(5):923–938.

4. Costa F, Hagan JE, Calcagno J, Kane M, Torgerson P, Martinez-Silveira MS, et al. Global morbidity and mortality of leptospirosis: a systematic review. PLoS Negl Trop Dis. 2015;9(9):e0003898.

5. Torgerson PR, Hagan JE, Costa F, Calcagno J, Kane M, Martinez-Silveira MS, et al. Global burden of leptospirosis: estimated in terms of disability adjusted life years. PLoS Negl Trop Dis. 2015;9(10):e0004122.

6. Karpagam KB, Ganesh B. Leptospirosis: a neglected tropical zoonotic infection of public health importance—an updated review. Eur J Clin Microbiol Infect Dis. 2020;39(5):835–846.

7. Lau CL, Smythe LD, Craig SB, Weinstein P. Climate change, flooding, urbanisation and leptospirosis: fuelling the fire? Trans R Soc Trop Med Hyg. 2010;104(10):631–638.

8. Phosri A. Effects of rainfall on human leptospirosis in Thailand: evidence of multi-province study using distributed lag non-linear model. Stoch Environ Res Risk Assess. 2022;36(12):4119–4132.

9. Matsushita N, Ng CFS, Kim Y, Suzuki M, Saito N, Ariyoshi K, et al. The non-linear and lagged short-term relationship between rainfall and leptospirosis and the intermediate role of floods in the Philippines. PLoS Negl Trop Dis. 2018;12(4):e0006331.

10. Douchet L, Menkes C, Herbreteau V, Larrieu J, Bador M, Goarant C, et al. Climate-driven models of leptospirosis dynamics in tropical islands from three oceanic basins. PLoS Negl Trop Dis. 2024;18(4):e0011717.

11. Ko AI, Galvão Reis M, Ribeiro Dourado CM, Johnson WD Jr, Riley LW. Urban epidemic of severe leptospirosis in Brazil. Lancet. 1999;354(9181):820–825.

12. Hagan JE, Moraga P, Costa F, Capian N, Ribeiro GS, Wunder EA Jr, et al. Spatiotemporal determinants of urban leptospirosis transmission: four-year prospective cohort study of slum residents in Brazil. PLoS Negl Trop Dis. 2016;10(1):e0004275.

13. Reis RB, Ribeiro GS, Felzemburgh RDM, Santana FS, Mohr S, Melendez AXTO, et al. Impact of environment and social gradient on Leptospira infection in urban slums. PLoS Negl Trop Dis. 2008;2(4):e228.

14. Felzemburgh RDM, Ribeiro GS, Costa F, Reis RB, Hagan JE, Melendez AXTO, et al. Prospective study of leptospirosis transmission in an urban slum community: role of poor environment in repeated exposures to the Leptospira agent. PLoS Negl Trop Dis. 2014;8(5):e2927.

15. Hacker KP, Sacramento GA, Cruz JS, de Oliveira D, Nery N, Lindow JC, et al. Influence of rainfall on Leptospira infection and disease in a tropical urban setting, Brazil. Emerg Infect Dis. 2020;26(2):311–314.

16. Sanders EJ, Rigau-Pérez JG, Smits HL, Deseda CC, Vorndam VA, Aye T, et al. Increase of leptospirosis in dengue-negative patients after a hurricane in Puerto Rico in 1996. Am J Trop Med Hyg. 1999;61(3):399–404.

17. Centers for Disease Control and Prevention. Initial public health laboratory response after Hurricane Maria — Puerto Rico, 2017. MMWR Morb Mortal Wkly Rep. 2018.

18. Jones FK, Medina AG, Ryff KR, Irizarry-Ramos J, Wong JM, O’Neill E. Leptospirosis outbreak in aftermath of Hurricane Fiona — Puerto Rico, 2022. MMWR Morb Mortal Wkly Rep. 2024;73(35):763–768.

19. Pan American Health Organization. Ready before the first case: tackling leptospirosis after Hurricane Melissa. Washington, DC: PAHO; 22 Nov 2025.

20. Martin BM, Zhang Z, Vernal S, Jian H, Nilles EJ, Furuya-Kanamori L, et al. Leptospirosis in the Caribbean Region between 2000 and 2022: a scoping review of morbidity and mortality. PLoS Negl Trop Dis. 2026.

21. Peters A, Vokaty A, Portch R, Gebre Y. Leptospirosis in the Caribbean: a literature review. Rev Panam Salud Publica. 2017;41:e166.

22. Gutiérrez JD, Martínez-Vega RA, Botello H, Ruiz-Herrera FJ, Arenas-López LC, Hernandez-Tellez KD. Environmental and socioeconomic determinants of leptospirosis incidence in Colombia. Cad Saude Publica. 2019;35(5):e00118417.

23. Martin BM, Sartorius B, Mayfield HJ, Cadavid Restrepo AM, Kiani B, Then Paulino CJ, et al. Geospatial analysis of leptospirosis clusters and risk factors in two provinces of the Dominican Republic. PLoS Negl Trop Dis. 2025;19(6):e0013103.

24. Martin BM, Sartorius B, Mayfield HJ, Cadavid Restrepo AM, Kiani B, Then Paulino CJ, et al. Quantifying spatial variation in environmental and sociodemographic drivers of leptospirosis in the Dominican Republic using a geographically weighted regression model. Sci Rep. 2025;15(1):27073.

25. Nilles EJ, Paulino CT, Galloway R, de St Aubin M, Mayfield HJ, Cadavid Restrepo A, et al. Seroepidemiology of human leptospirosis in the Dominican Republic: a multistage cluster survey, 2021. PLoS Negl Trop Dis. 2024;18(12):e0012463.

26. Chadsuthi S, Modchang C, Lenbury Y, Iamsirithaworn S, Triampo W. Modeling seasonal leptospirosis transmission and its association with rainfall and temperature in Thailand using time-series and ARIMAX analyses. Asian Pac J Trop Dis. 2012;2(6):539–546.

27. Semenza JC, Ko AI. Waterborne diseases that are sensitive to climate variability and climate change. N Engl J Med. 2023;389(23):2175–2187.

28. Lotto Batista M, Rees EM, Gómez A, López S, Castell S, Kucharski AJ, et al. Towards a leptospirosis early warning system in northeastern Argentina. J R Soc Interface. 2023;20(202):20220896.

29. Durski KN, Jancloes M, Chowdhary T, Bertherat E. A global, multi-disciplinary, multi-sectorial initiative to combat leptospirosis: Global Leptospirosis Environmental Action Network (GLEAN). Int J Environ Res Public Health. 2014;11(6):6000–6008.

30. Luangnara A, Towachiraporn S, Wiwatkunupakarn N, Thongwitokomarn H. Impact of flooding events on waterborne and vector-borne infections: a systematic review. BMC Infect Dis. 2026;26(1):112.

31. Cunha M, Costa F, Ribeiro GS, Carvalho MS, Reis RB, Nery N, et al. Rainfall and other meteorological factors as drivers of urban transmission of leptospirosis. PLoS Negl Trop Dis. 2022;16(4):e0010382.

32. Alcántara LV, Sánchez JJ, De Luna D, Aleuy OA, Miller BL, Mace CR, et al. Epidemiology, temporal and seasonal trends, and geographic distribution of leptospirosis in the Dominican Republic, 2012–2026. Manuscript in preparation.

33. Gasparrini A, Armstrong B, Kenward MG. Distributed lag non-linear models. Stat Med. 2010;29(21):2224–2234.

34. Gasparrini A. Distributed lag linear and non-linear models in R: the package dlnm. J Stat Softw. 2011;43(8):1–20.

35. Ver Hoef JM, Boveng PL. Quasi-Poisson vs. negative binomial regression: how should we model overdispersed count data? Ecology. 2007;88(11):2766–2772.

36. Greenland S, Drescher K. Maximum likelihood estimation of the attributable fraction from logistic models. Biometrics. 1993;49(3):865–872.

